# The same practices, year after year: a national observational study of higher-risk antibiotic prescribing in English general practice, 2023–2026

**DOI:** 10.64898/2026.11.18.26368426

**Authors:** Ashish Agarwal, Aashish Gill

## Abstract

**Background:** There is increasing antibiotic resistance in England. National targets have been set to increase the proportion of antibiotics prescribed in the lower risk Access category. The proportion of higher risk antibiotics within the Watch+Reserve category prescribed in each general practice is publicly available. It is unclear whether a high proportion of Watch+Reserve antibiotics in a practice is due to random variation, patient population or a persistent trend.

**Methods:** Observational study using NHS open prescribing data from 5,791 general practices in England (26.0 million items, September 2025 to August 2026, and the two previous years). Watch+Reserve proportions were compared between practices using over-dispersion adjusted funnel plots with 99.8% control limits.

Practices identified as outliers were tested using subsequent data. A quasi-binomial model was fitted adjusting for age, deprivation, practice size and 7 disease registers.

**Results:** 117 practices exceeded the 99.8% control limit (approximately 6 expected by chance). 106 practices were included in a shortlist. Of those in the shortlist in 2024/25, 61% (95% CI 52–70%) exceeded the control limit in 2025/26 compared to 1.0% of all other practices. 36 practices were shortlisted in all 3 years. Adjusting for age, deprivation, practice size and disease registers explained 4.1% of variability. Following adjustment, 101 of 106 practices remained outliers. Overall, shortlisted practices did not prescribe more antibiotics than other practices (median 422 vs 438 items per 1,000 patients per year), but did prescribe fewer Access antibiotics and more Watch+Reserve antibiotics. 10% or fewer of excess Watch+Reserve antibiotics in England were prescribed in shortlisted practices.

**Conclusions:** A small number of practices show persistently high prescribing of higher risk antibiotics, which cannot be explained by recorded need. This provides integrated care boards (ICBs) with a reliable list of practices to review. It was not possible to investigate penicillin allergy. To meet national Access targets, interventions will be needed in all practices, particularly reviewing the use of clarithromycin, co-amoxiclav and azithromycin.

## Introduction

There were 21,129 cases of antibiotic resistant bacteraemia in England in 2025 (an increase of 16.8% compared to 2019), and general practice accounted for 65% of antibiotics prescribed within the NHS (UKHSA, 2026). It is important to consider not only how many antibiotics are prescribed from a practice, but also which antibiotics are prescribed.

Antibiotics can be categorised into three groups based on their risk of resistance development: Access, Watch and Reserve (UKHSA, 2025). The UK National Action Plan aims to reduce the use of antibiotics in non-Access groups to have 70% of total antibiotic use in the Access group by 2029. Within primary care, NHS England has set a specific target of 73% (HM Government, 2024; NHS England, n.d.). Individual practice data on the proportion of Watch and Reserve antibiotics prescribed is available, however it is unclear if a high proportion is due to random variation, patient population, or practice routine.

In 2019, it was estimated that 1.27 million global deaths were a result of antimicrobial resistance in bacteria (Antimicrobial Resistance Collaborators, 2022). As certain antibiotics are more likely to contribute to resistance development than others, national targets have been set to control both the quantity and type of antibiotics prescribed. In this study, the proportion of Watch+Reserve antibiotics has been selected to indicate the type of antibiotics being prescribed.

Most literature relates to volume. Prescribing in practices with a higher proportion of elderly people, those in more deprived areas and in rural areas was higher (Devine, O’Kane and Bucholc, 2021). In a wider review of 23 studies in the UK, 21 showed increased prescribing in more deprived areas (Bell et al., 2026). However, neither study looked at the proportion of higher risk antibiotics or whether a practice’s position lasts. The most recent literature to relate to this question considers area levels or overall volume. A number of commissioning areas had the highest prescribing in 2021, 2022, 2023 and 2024 (Lee, Lee and Lee, 2026 [preprint]), however this was not tested against chance. In another study, variation in prescribing was split between patients, prescribers and practices in the UK, but no outlier practices were identified (Nguyen et al., 2026 [preprint]).

Watch and Reserve antibiotics may be prescribed at higher proportions in certain practices for clinical reasons. Lymecycline (Watch antibiotic, mainly prescribed for acne) is often prescribed for long durations (Bhate et al., 2023). Clarithromycin (most commonly prescribed Watch antibiotic) is a common alternative when a penicillin allergy is recorded. However, many of these recorded allergies are incorrect. In the ALABAMA trial, after assessment, penicillin was prescribed for first line infections in 18% of patients with a recorded allergy, compared to 3% without assessment (Sandoe et al., 2025).

Feedback has a small effect on reducing prescribing. Sending feedback letters to high prescribing practices in England reduced prescribing volumes by 3.3% over 6 months (Hallsworth et al., 2016). Peer comparison letters to practices in Ontario reduced prescribing by approximately 4% (Schwartz et al., 2021). These interventions targeted overall volume. Targeted feedback to reduce broad spectrum antibiotic use had no effect (Gold et al., 2022).

Monthly data is available for each practice on OpenPrescribing (Bennett Institute for Applied Data Science, 2026), however it does not test whether a high proportion is due to chance.

A non-systematic search was conducted in November 2026. No literature was found which looked at outlier prescribing in practices for this proportion, with analysis in subsequent years, and adjustment for known disease to quantify over prescription. Therefore this study makes a contribution in this area. However, as the search was non-systematic, this conclusion is limited to what the search found.

To help ICB medicines optimisation teams decide which practices to prioritise, this study aimed to answer the following questions:

1. Which practices have an unusually high proportion of Watch and Reserve antibiotics prescribed, after adjusting for random variation?
2. Have these practices had unusually high proportions in subsequent years?
3. Is this proportion explained by demographics (age, deprivation) and recorded illness?
4. How much of the excess Watch and Reserve prescribing in England is held by these practices?

## Methods

Items refer to prescriptions. ICB (integrated care board) refers to the local NHS body which the practice belongs to. This is an observational study of open, aggregated NHS data, reported following the STROBE guidelines (Supplementary file 1).

Antibiotic prescribing data was extracted from the NHSBSA English Prescribing Dataset using SQL through its API. Total antibiotic prescriptions were calculated by practice and month between September 2023 and August 2026. The API output for July 2026 was identical to the full dataset (2,361,207 prescriptions), before 4,505 items not linked to a practice were removed. Data was linked to practice demographics (patient list size, age banding), deprivation (IMD 2025) and 7 QOF disease registers in 2025/26. After removing mismatched data, practices with less than 1,000 patients, partial data and 59 anomalous patient lists, 5,791 practices remained with 26.0 million coded prescriptions. Of 81 substances, 14 not included in the UK-AWaRe list (mainly for TB and leprosy) were excluded from the proportion.

Watch+Reserve proportion: proportion of Watch and Reserve prescriptions within coded prescriptions. A funnel plot was created to account for overdispersion (Spiegelhalter, 2005). The limits were widened to account for natural variation in proportions across practices. The z-score for each practice was calculated using 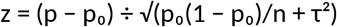, where p is the proportion, p_0_ is the proportion in England, n is the number of prescriptions, and τ is the natural spread between practices (2.2 percentage points). If z > 3.09, the practice is outside of the 99.8% limit. Practices were shortlisted if they were beyond the 99.8% limit, in the highest 20% for proportion in 9 or more of 12 months, and still in the highest 20% in 9 or more of 12 months when excluding lymecycline. The monthly criteria were chosen so that a practice had to be high for most of the year, and could not be flagged by a seasonal peak or by acne prescribing alone. These cut-offs are a judgement rather than an optimised value, so the number of practices identified under other funnel thresholds is also reported.

Robustness of flags was assessed by applying the screening method independently to the first half of 2025/26 and previous years, and tracking the flagged practices. A quasi-binomial model was used to predict the expected proportion for each practice based on disease prevalence, age, deprivation and practice size. The funnel plot was recreated using this expected value. Practices were grouped by ICB in August 2026. Mergers meant that there were 36 ICBs in 2026, compared to 42. All key results have been calculated using Excel, with the three-year and case-mix models in Python, and validated using an independent calculation in Python. Confidence intervals for proportions were calculated using Wilson intervals. All data is open and aggregated. No patient level information was used, and practices are not named.

## Results

### 106 practices stand out beyond chance

A plain funnel plot (allowing only for chance) has 1,405 practices above its 99.8% limit, which is far too many given that practices are not all the same. After adjustment 117 are still above the limit (about 6 would be expected by chance alone), so a number of those may still be false positives (Figure 1). Of those 117, 106 meet all three screening criteria and constitute the review shortlist.

**Figure 1.**
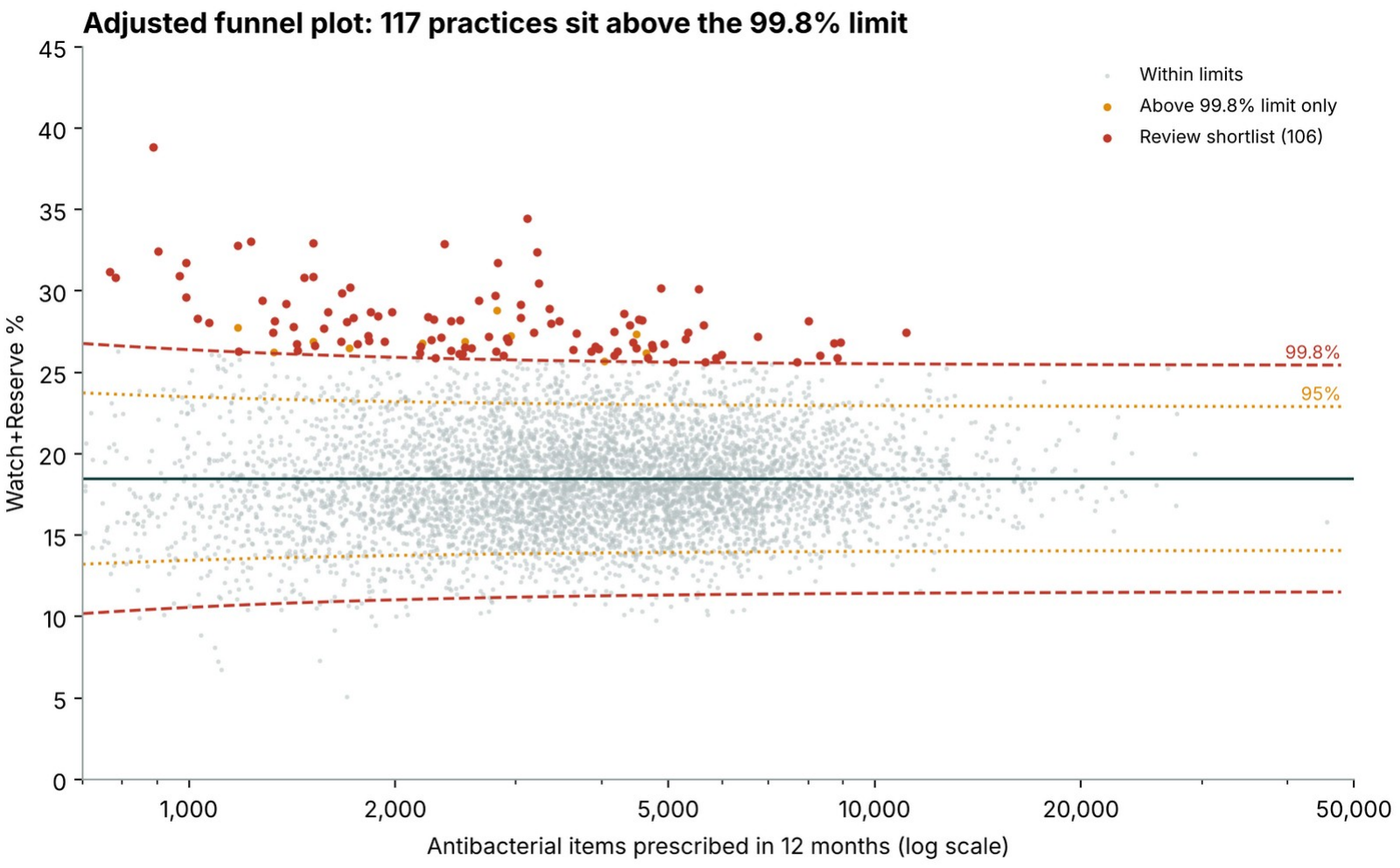
Adjusted funnel plot of Watch+Reserve share against items prescribed, with 95% and 99.8% limits. Source: authors’ analysis of NHS Business Services Authority (2026).

Examination of out-of-sample months (March to August) indicates that 56% (95% CI 46–65%) of practices identified as outliers (September to February) remain above the 99.8% limit, compared to 0.9% of unflagged practices. Their mean proportion did not change substantially (28.1% to 27.6%), compared to 18.9% for England.

The total number of practices identified depends on the threshold. At z > 2.58, 182 practices were identified and at z > 3.5, 72 practices. When comparisons are made only within the practice’s ICB, 93 of the 106 practices remain identified. Therefore, the list is mostly driven by individual practices, not regional effects. The distribution is still uneven, with ten ICBs having no shortlisted practices, but 17 practices in North East and North Cumbria. Practices have been ranked according to adjusted z-scores. Ranking by number of excess items means larger practices are more likely to be at the top of the list. Only 4 of the top 15 practices match.

### The same practices, year after year

A more stringent test is a whole separate year. Of those practices shortlisted in 2024/25, 61% (95% CI 52– 70%) had results above the 99.8% limit in 2025/26, compared to 1.0% of non-shortlisted practices. There was strong correlation between the proportions across the two years (r = 0.87) (Figure 2). Of 111 practices shortlisted in 2023/24, 44% (35–53%) were above the limit in 2025/26, compared to 1.2%.

**Figure 2.**
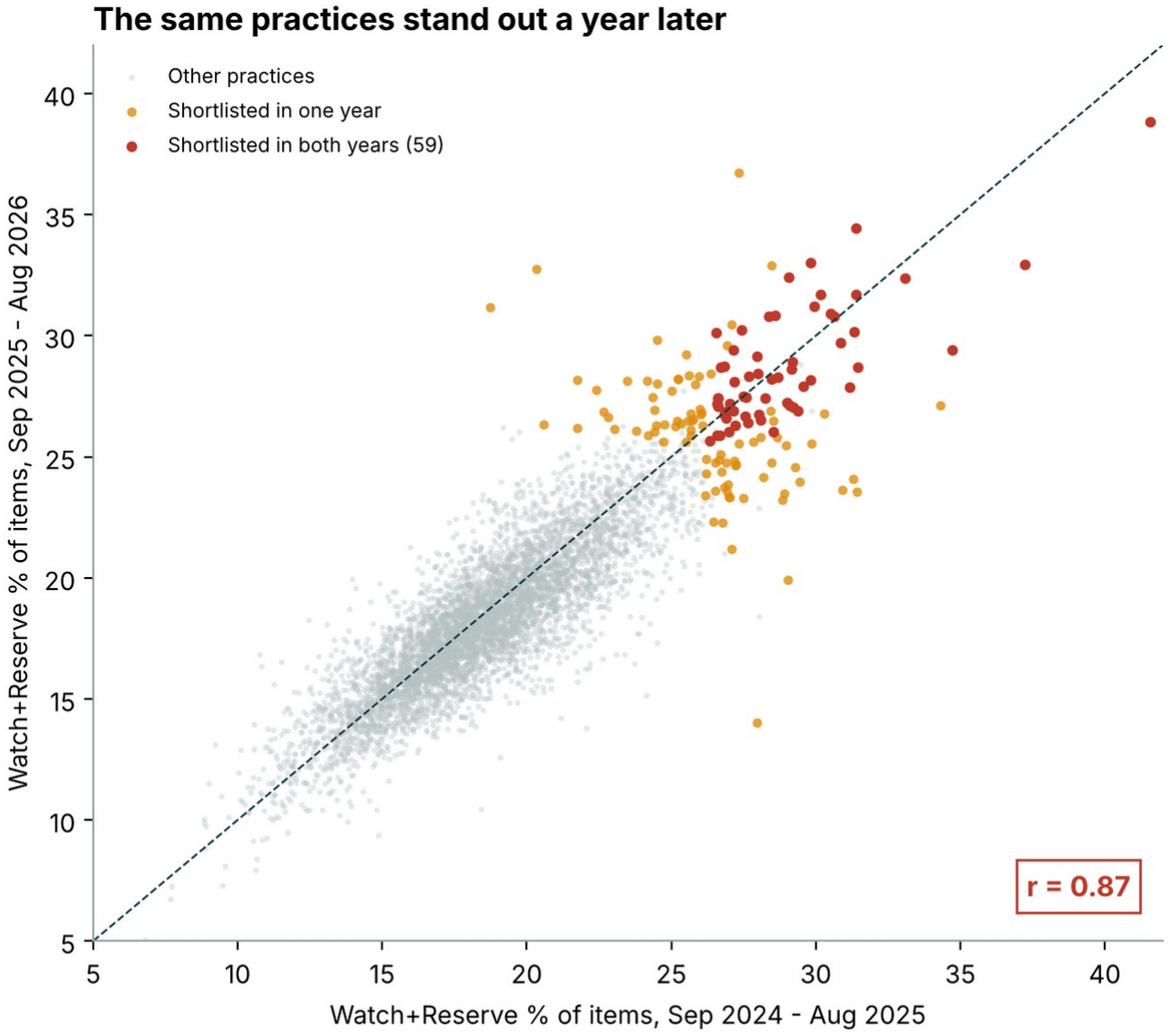
Watch+Reserve share in September 2024–August 2025 against September 2025–August 2026, each practice. Red: shortlisted in both years; amber: shortlisted in one year. Source: authors’ analysis of NHS Business Services Authority (2026).

There appears to be some decay in the signal over time. However, the confidence intervals of the two periods nearly meet, so the difference is uncertain. Some decline is expected from regression to the mean, as practices flagged partly by chance in one year will move towards the average in the next. That 61% of shortlisted practices remained above the limit, compared with 1.0% of other practices, indicates that most of the signal is not explained by this. 36 practices were shortlisted in all three years, and 88 were shortlisted in at least two years.

There are two further considerations. First, the mean proportion of flagged practices reduced slightly between years, to 26.8% from 28.6%. This suggests that some practices may have been flagged by chance in single years, and therefore practices flagged in two years provide stronger evidence than single year flags. Second, practices with a consistently low proportion are less stable than practices with a consistently high proportion. Only 19 of the 48 practices that were significantly low in 2025/26 had also been significantly low in the previous year. These should therefore not yet be considered best practice.

### Patient need does not explain it

One concern is that high-proportion practices may simply have sicker patients. However, only a small amount of variance in proportion can be explained by the health needs of the population. Age, deprivation and practice size explain 1.5% of variation. Adding 7 disease registers increases the variance explained by 2.5 percentage points (to 4.1%). Practices in the shortlist do not have higher levels of disease. The median prevalence of COPD is 1.82% compared to 1.91%.

Only 5 of the 106 shortlisted practices fall back within the 99.8% limit when the funnel plot is adjusted for each practice’s expected proportion (Figure 3). This is a weak test, as the adjustment made few changes to each practice’s score, and QOF does not include registers for penicillin allergy, bronchiectasis or acne.

**Figure 3.**
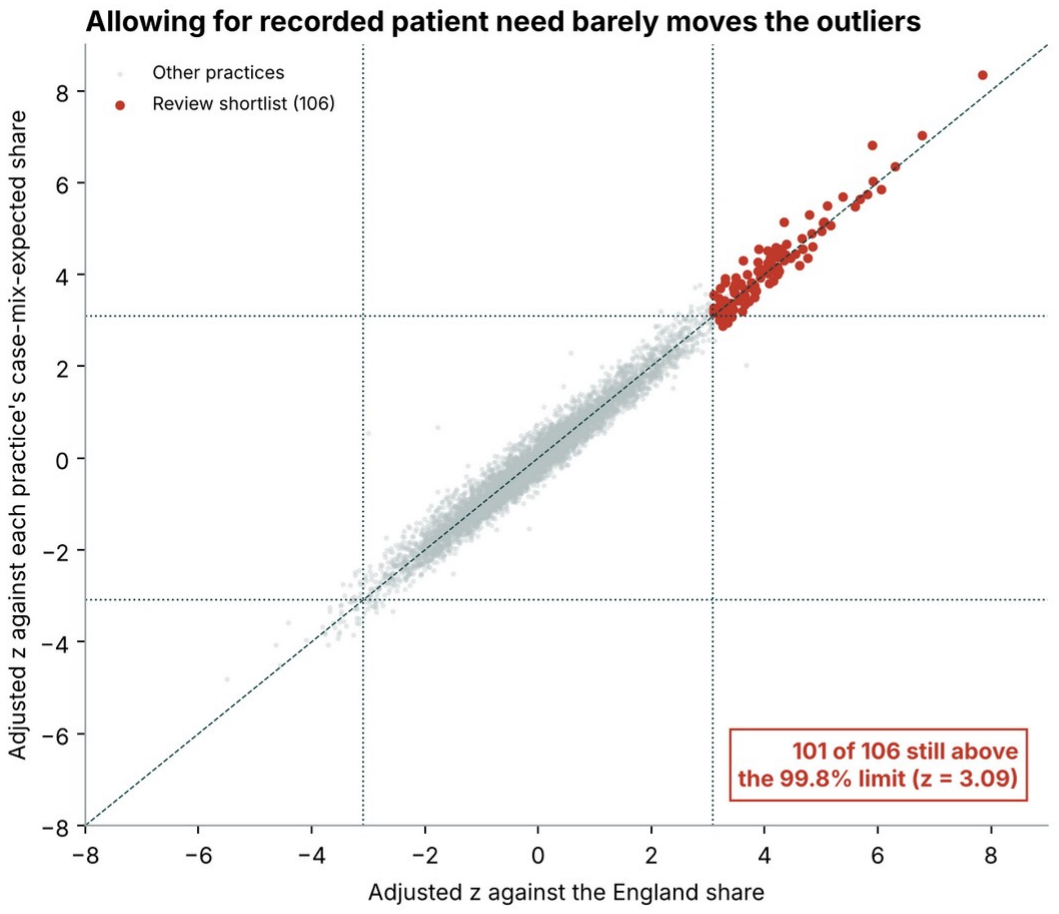
Adjusted funnel z against the England share (x) and against each practice’s case-mix-expected share (y). Dotted lines: 99.8% limits. Source: authors’ analysis of NHS Business Services Authority (2026) and NHS England Digital (2026).

Therefore, these registers (but not patients in general) can be ruled out.

Age and deprivation affect volume, not the proportion. Older lists prescribe more (R^2^ = 0.34), but the proportion remains largely unchanged (r = 0.04). After adjusting for age and practice size, volume prescribed increased with increasing deprivation (3.4 items per 1,000 patients for every unit increase in IMD). However, there was only a weak association between deprivation and proportion prescribed, which was not robust after accounting for clustering of errors within ICBs (p = 0.10). Deprivation is confounded by patient age in the data: practices with more deprived populations had younger patients (r = −0.46), who were prescribed fewer antibiotics. Averaging across the ICB explains 13.2% of the variation in proportion prescribed. This is similar to the variation in total prescribing explained by area membership (12.6%) found by Curtis et al. (2019). However, adding age, deprivation and practice size adds less than one percentage point to this. This suggests that most variation in proportion comes from differences between practices, in prescribing patterns or in unmeasured patient need.

### Swapping, not over-prescribing

Although both higher Watch and Reserve prescribing and lower Access prescribing lead to a high Watch and Reserve proportion, shortlisted practices do not prescribe more overall. They prescribed a median of 422 items (per 1,000 patients per year) compared to 438 items in other practices (Table 1). They do however prescribe less Access (294 vs. 343) and more Watch and Reserve (118 vs. 78) than other practices, which is the substitution stewardship teams look out for. However, this does not occur in all 106 shortlisted practices. 32 shortlisted practices had a high proportion partly due to low Access prescribing. 559 practices with high Watch and Reserve prescribing were not identified, because they also prescribe a lot of Access.

**Table 1.** Median items per 1,000 patients a year.

|  | **Access** | **Watch and Reserve** | **Total** |
|---|---|---|---|
| The 106 shortlisted practices | 294 | 118 | 422 |
| All other practices | 343 | 78 | 438 |

### The outliers hold a tenth of the problem or less

If all practices on the shortlist prescribed at the England proportion, about 31,000 Watch and Reserve items a year would become Access. This is 10% of the approximately 299,000 items above the England level, and 4% when the best 10% of practices is used as the benchmark (Figure 4). If all 299,000 items were switched, England’s Access proportion would increase by 1.2 percentage points, whereas switching only the shortlist would increase it by 0.12 points. These figures are item-level based, and some practices have higher prescribing than England for good clinical reasons. Regardless of the benchmark, the majority of excess prescribing is found in approximately 2,500 practices that are just above the England level.

**Figure 4.**
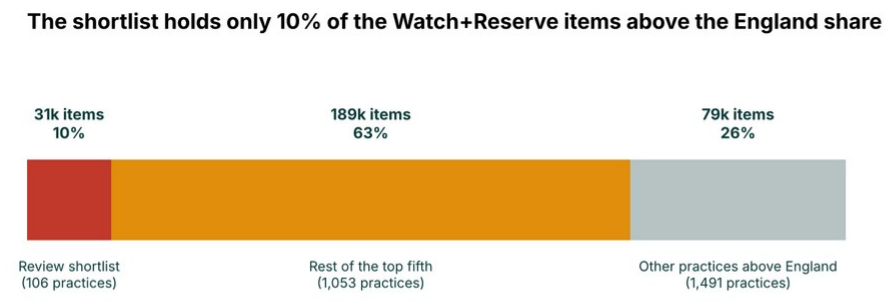
Watch+Reserve items above the England share, by group of practices, 12 months. Source: authors’ analysis of NHS Business Services Authority (2026).

The 48 practices significantly below the lower limit show that a low proportion does not necessarily mean under-prescribing. These practices have similar Access prescribing to other practices (340 vs. 342 items per 1,000 patients) but about half the Watch and Reserve prescribing (41 vs. 79).

### What drives the proportion, and why a year matters

Clarithromycin (29.6%), co-amoxiclav (20.2%) and azithromycin (18.7%) account for 68.5% of Watch items. Lymecycline accounts for a further 15.4% (Figure 5B). If lymecycline is excluded from the analysis, the national proportion drops by 2.4 percentage points, and 27% of the persistently high practices change. If we consider the dose prescribed rather than items, the national proportion increases to 22.9% due to long durations of acne treatment. 62 of the 106 practices remain high for 9 months or more. The remaining 44 practices still have a median proportion of 26.8%, so using the dose would mean a different ranking for these practices, but they would not be cleared. Other analysis choices change the results less (Table 2).

**Table 2.** Headline results under different choices. “High in 9+ months” means in the monthly top fifth in at least 9 of 12 months.

| Version | England Watch+Reserve % | Practices high in 9+ months | Same practices as the main list |
|---|---|---|---|
| Main analysis (items, 5,791 practices) | 18.5 | 515 | 515 |
| Without lymecycline | 16.1 | 496 | 375 |
| OpenPrescribing denominator (Access + Watch + Reserve) | 19.1 | 522 | 458 |
| By dose (ADQ) instead of items | 22.9 | 360 | 198 |
| Keeping the 59 atypical practices | 18.5 | 516 | 509 |
| Each practice compared with its own ICB | 18.5 | 120 shortlisted | 93 of the 106 |

**Figure 5.**
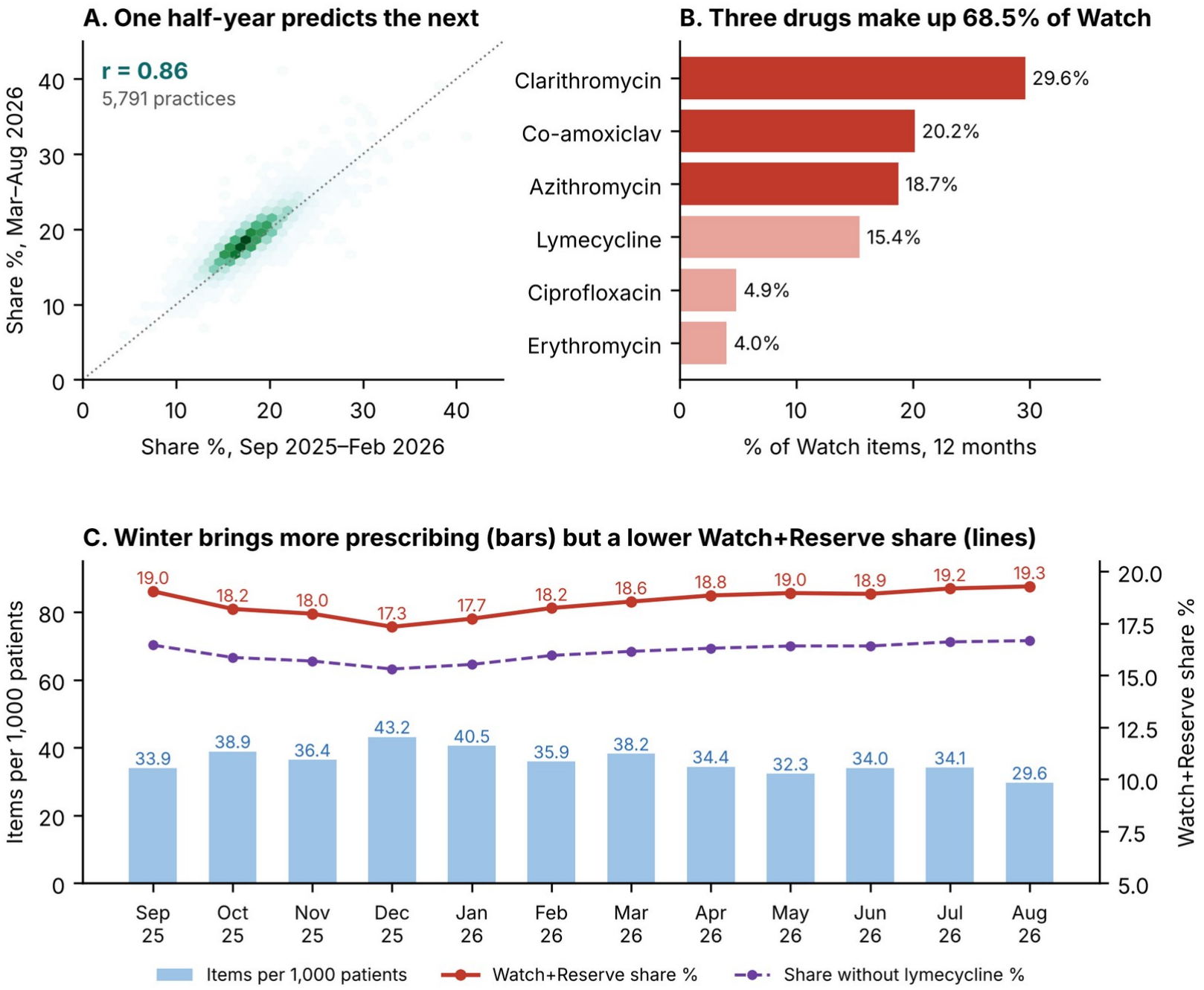
(A) Each practice’s share in the two halves of the year; (B) the six most-prescribed Watch antibiotics; (C) items per 1,000 patients and Watch+Reserve share by month. Source: authors’ analysis of NHS Business Services Authority (2026) and NHS England (2026).

Yearly prescribing proportions correlate well between the two half-years (r = 0.86) (Figure 5A). Single month proportions are misleading. About 36% of the differences between practices in a single month are due to chance, compared to 5% over 12 months. Only 7 of the top 15 practices in July matched the top 15 practices over 12 months. There is clear seasonality. Although prescribing levels were highest in December, Watch and Reserve prescribing as a proportion of total prescribing was lowest (17.3%) in December, as increased winter prescribing was largely Access. As such, summertime proportions overstate the yearly proportion in a practice (Figure 5C).

## Discussion

The Watch+Reserve proportion is a stable characteristic of a practice. Correlation between half-year periods was high (r = 0.86), and flags held in later months and in later years. As seen in Table 3, this research supports previous findings in the UK relating to age, deprivation and volume. However, the stability of this proportion and its resistance to adjustment for case mix are new findings.

**Table 3.** Findings compared with published research.

| Finding in this report | Published evidence | Interpretation |
|---|---|---|
| **Established findings** |  |  |
| Older lists prescribe more (R<sup>2</sup> = 0.34) | More prescribing where more patients are 65+ (Devine, O’Kane and Bucholc, 2021) | Age goes with how much is prescribed, not which drugs |
| Practice size barely affects volume (r = −0.10) | No effect of practice size (Devine, O’Kane and Bucholc, 2021) | Size is no reason to flag a practice |
| More deprived practices prescribe more once age is held constant | 21 of 23 UK studies link deprivation with more prescribing (Bell et al., 2026) | Matches, but the link only shows once younger lists are allowed for |
| ICB average explains 13.2% of the variation in share | Area membership explained 12.6% of practice prescribing (Curtis et al., 2019) | Area matters about as much as before; most differences lie within areas |
| Lymecycline changes 27% of the persistent list | 44.5% of new acne patients had long-term oral antibiotics (Bhate et al., 2023) | Long acne courses help explain why removing it changes the list |
| **Additional contributions** |  |  |
| Registers, age and deprivation explain 4.1% of the share; 101 of 106 outliers remain | These explained 7.5–9.1% of total prescribing; a fifth of high prescribers reclassified (Hope et al., 2018) | Little explained in both studies; new is that it clears almost no outliers |
| Deprivation and share: r = −0.12, not robust once ICB is allowed for | Earlier studies examine volume, not the share | Small, and not robust once ICB is allowed for |
| Share is stable within a year (r = 0.86); single months are noisy | No comparison found for the Watch+Reserve share in England | A full year gives a reliable ranking; one month does not |
| Flags hold a year later (61% against 1.0%) and two years later (44% against 1.2%) | No comparison found for the Watch+Reserve share in England | The list reflects lasting patterns, not chance |

A further alternative explanation for the findings may be the presence of penicillin allergy labels. If there are more of these labels in the selected practices, then in part the high proportion could be due to records rather than choices. The presence of these labels in practices can be identified at local level within the ICB. Similarly, the presence of bronchiectasis and long-term azithromycin for COPD, which are appropriate indications for prescribing Watch, cannot be seen in the published data. This limits the conclusions we can make on persistence. Practices with many allergy labels would be expected to stand out each year in the same way as practices with fixed prescribing habits. Both situations would result in the same change from Access to Watch. While we can see that this pattern is real and consistent, we cannot currently determine how much of it is down to prescribing habits.

However, due to the low amount of excess (10% or less) in these outliers, reviewing these practices is a cheap first step but cannot on its own move the national figure. The larger impact will come from improving the use of the three drugs which make up the majority of Watch prescribing across England. Nationally, the Access rate in primary care in 2025 is 68.9%, which is below the national 70% target, with NHS England asking primary care for 73% by 2029 (UKHSA, 2026; NHS England, n.d.). This was measured in defined daily doses (DDDs), whereas in this study the Access rate is 78.0% by items, meaning only comparison between practices can be made, and not progress towards the target.

Evidence shows that feedback letters were only slightly effective at reducing overall volume, and in the one trial aimed at the proportion of broad-spectrum antibiotics, there was no effect (Gold et al., 2022). A review of the mix is more likely to be effective if it is a discussion on particular drugs and patients rather than a letter.

### Strengths and limitations

The main strengths are national coverage of English general practice, three years of data, and testing of flagged practices on data that was not used to flag them. The main limitations are:

1. **No diagnosis information**. Some practices in the shortlist may be high for appropriate clinical reasons (see Results and Discussion). Being flagged starts a review; it does not indicate a judgement.
2. **Items, not doses**. The national target is measured in doses. When measured by dose, the shortlist is re-ranked but not cleared (Table 2), but progress towards the target cannot be measured with this measure.
3. **Repeats**. As items are not independent decisions, the noise figures may be underestimates. Additional spread is accounted for by the τ adjustment within the funnel.
4. **Proportion**. This can be high because of low use of Access. Practices with high Watch and Reserve and high Access prescribing may be missed (see Results).
5. **Area level variables**. Deprivation is measured for areas rather than individuals. Age bands were only obtained from July 2026, and rurality and care homes were not investigated.
6. **3 years**. The year 2022/23 was excluded due to an outbreak of Group A streptococcus which increased Watch prescribing (Cunningham et al., 2024). Whether flagged practices change following review has not been investigated.

### Implications for practice

For ICB medicines optimisation teams, the findings suggest the following actions:

1. **Consider the 36 practices identified across all 3 years first**, then consider the rest of the 106 practices, prioritising those with high Watch and Reserve items per 1,000 patients.
2. **Examine the penicillin allergy data**. For practices on the shortlist, compare the proportion of patients with a penicillin allergy label to the average for the ICB. If practices have a high proportion, consider offering an allergy assessment. This addresses the most likely confounder. If patients were mislabelled with an allergy, they can be prescribed first line penicillins.
3. **Differentiate between acne prescribing and acute prescribing**. The length of lymecycline courses should be compared separately to the 24-week duration target set by NHS England. This is to avoid masking issues in acute prescribing.
4. **Prioritise reviewing prescribing of clarithromycin, co-amoxiclav and azithromycin**. These 3 drugs account for more than two thirds of Watch items, therefore providing guidance on appropriate use of these drugs will address most Watch prescribing in a practice.
5. **Only consider 12 month periods**. Using single month periods would be too volatile and affected by seasonality (see Results).
6. **Consider actions beyond this list**. Consider providing guidance for all practices in the ICB on the key Watch drugs. After 1 year, the adjusted funnel plot can be repeated to see if any changes are seen within shortlisted practices.
7. **Consider expanding the analysis**. Include an additional measure using defined daily doses to monitor progress towards the national target. If available, combine the penicillin allergy and bronchiectasis data within the ICB. Follow up the highlighted practices for an additional year.

## Conclusion

Practices with a high Watch+Reserve proportion were identified. These practices show a sustained, genuine pattern, are not explained by recorded need, and represent only a small proportion of the national excess. This shortlist is therefore a good starting point, however the reviews should include consideration of reasons for variation in prescribing that cannot be captured in the data, such as allergy to penicillin. Ranking over a full year, validating with subsequent data, and considering case mix means that this shortlist can be relied upon by the ICB, and the same method could be used for other prescribing measures. However, meeting the national target relies on all practices performing well.

## Supporting information

Supplementary file 1: STROBE checklist

## Data Availability

All data used in this study are publicly available from the NHS Business Services Authority (English Prescribing Dataset), NHS England (practice list sizes and Quality and Outcomes Framework), the Ministry of Housing, Communities and Local Government (English indices of deprivation 2025) and the UK Health Security Agency (UK-AWaRe classification). The analysis code, workbook and outputs are available on GitHub (https://github.com/gagarwalashish1-hue/antibiotic-stewardship-england). An interactive dashboard of the results is available on Tableau Public (https://public.tableau.com/app/profile/ashish.agarwal6382/viz/AntibioticStewardshipinEnglishGPPractices/Dashboard).

https://public.tableau.com/app/profile/ashish.agarwal6382/viz/AntibioticStewardshipinEnglishGPPractices/Dashboard

## Declarations

### Ethics approval

Not required. The study used only open, aggregated data published at practice level. No patient-level data were used, and practices are not named.

### Data availability

All data are publicly available from the NHS Business Services Authority (English Prescribing Dataset), NHS England (practice list sizes and Quality and Outcomes Framework), the Ministry of Housing, Communities and Local Government (English indices of deprivation 2025) and the UK Health Security Agency (UK-AWaRe classification).

### Code availability

The SQL, Python and Excel workbook used for the analysis are available at https://github.com/gagarwalashish1-hue/antibiotic-stewardship-england. An interactive dashboard is available on Tableau Public.

### Competing interests

The authors declare no competing interests.

### Funding

This research received no specific funding.

### Author contributions

Ashish Agarwal conceived and designed the study, extracted the data, carried out the analysis and wrote the manuscript. Aashish Gill created the visualisations and contributed to the interpretation of the results. Both authors reviewed and approved the final version.

### Supplementary material

Supplementary file 1: STROBE checklist. Additional figures and analyses are available in the project repository (see Code availability).

## Data licence and acknowledgements

Contains public sector information licensed under the Open Government Licence v3.0. Prescribing data: NHSBSA Copyright 2023-2026 (English Prescribing Dataset). Practice list sizes and QOF disease registers: NHS England. English indices of deprivation 2025: Ministry of Housing, Communities and Local Government. UK-AWaRe classification: UK Health Security Agency. National comparison figures: OpenPrescribing.net, Bennett Institute for Applied Data Science, University of Oxford.

## Notes

### Competing Interest Statement

The authors have declared no competing interest.

### Author Declarations

All source data are openly available to the public, with no application or registration required. Prescribing data: NHS Business Services Authority, English Prescribing Dataset (https://opendata.nhsbsa.net/dataset/english-prescribing-dataset-epd-with-snomed-code). Practice list sizes and age bands: NHS England Digital, Patients Registered at a GP Practice (https://digital.nhs.uk/data-and-information/publications/statistical/patients-registered-at-a-gp-practice). Disease registers: NHS England Digital, Quality and Outcomes Framework 2025-26 (https://digital.nhs.uk/data-and-information/publications/statistical/quality-and-outcomes-framework-achievement-prevalence-and-exceptions-data/2025-26). Deprivation: Ministry of Housing, Communities and Local Government, English indices of deprivation 2025 (https://www.gov.uk/government/statistics/english-indices-of-deprivation-2025). Antibiotic classification: UK Health Security Agency, UK-AWaRe classification (https://www.gov.uk/government/publications/uk-aware-antibiotic-classification). All data are aggregated at practice level; no individual patient data were used.

