## Supplementary file 1: STROBE checklist for "The same practices, year after year: a national observational study of higher-risk antibiotic prescribing in English general practice, 2023–2026"

### Supplementary file 1: STROBE checklist (cross-sectional / observational study)

*The same practices, year after year: a national observational study of higher-risk antibiotic prescribing in English general practice, 2023–2026*

| Item | No. | Recommendation (summary) | Where reported |
|---|---|---|---|
| Title and abstract | 1 | Indicate the design; give an informative, balanced abstract | Title; Abstract |
| Background/rationale | 2 | Explain the scientific background and rationale | Introduction, paragraphs 1–6 |
| Objectives | 3 | State specific objectives | Introduction, four study questions |
| Study design | 4 | Present key elements of study design early | Methods, paragraph 1 |
| Setting | 5 | Describe setting, locations, periods of data collection | Methods, paragraph 2 (England, Sep 2023 to Aug 2026) |
| Participants | 6 | Eligibility criteria and selection of participants | Methods, paragraph 2 (practice exclusions) |
| Variables | 7 | Define outcomes, exposures, confounders | Methods, paragraphs 2–3 (Watch+Reserve proportion; age, deprivation, size, QOF registers) |
| Data sources/measurement | 8 | Sources of data and methods of measurement | Methods, paragraph 2; Declarations (data availability) |
| Bias | 9 | Efforts to address potential sources of bias | Methods, paragraphs 3–4 (overdispersion, out-of-sample validation, case-mix model) |
| Study size | 10 | Explain how the study size was arrived at | Methods, paragraph 2 (5,791 practices after exclusions) |
| Quantitative variables | 11 | How quantitative variables were handled | Methods, paragraph 3 (z-score formula, limits) |
| Statistical methods | 12 | Describe all statistical methods, missing data, sensitivity analyses | Methods, paragraphs 3–4; Results, Table 2 (sensitivity analyses) |
| Participants (results) | 13 | Numbers at each stage | Methods, paragraph 2; Results, first subsection |
| Descriptive data | 14 | Characteristics of participants | Results; Table 1; project repository |
| Outcome data | 15 | Numbers of outcome events or summary measures | Results; Figures 1–5 |
| Main results | 16 | Unadjusted and adjusted estimates with precision | Results (95% confidence intervals; case-mix adjusted |

| Item | No. | Recommendation (summary) | Where reported |
|---|---|---|---|
| | | | funnel plot, Figure 3) |
| Other analyses | 17 | Subgroup, interaction and sensitivity analyses | Results, Table 2; project repository |
| Key results | 18 | Summarise key results with reference to objectives | Discussion, paragraph 1; Conclusion |
| Limitations | 19 | Discuss limitations, sources of bias and imprecision | Discussion, Strengths and limitations |
| Interpretation | 20 | Cautious overall interpretation | Discussion, paragraphs 2–4; Table 3 |
| Generalisability | 21 | External validity of the results | Discussion, Strengths and limitations (items 2 and 5) |
| Funding | 22 | Source of funding and role of funders | Declarations (no specific funding) |
